# The relationships between impulsivity and mood in bipolar disorder: An ecological momentary assessment study

**DOI:** 10.1101/2024.11.21.24317705

**Authors:** Almudena Ramírez-Martín, Lea Sirignano, Jerome C. Foo, Fabian Streit, Josef Frank, Stephanie H. Witt, Marcella Rietschel, Fermin Mayoral-Cleries, Berta Moreno-Küstner, Jose Guzmán-Parra

## Abstract

**Background:** Impulsivity is a key feature of bipolar disorder (BD) associated with various negative outcomes. Recent use of ecological momentary assessment (EMA) has allowed for nuanced examination of the mechanisms of mood and impulsivity dysregulation. However, few existing studies have used an ecological momentary assessment of impulsivity in multiplex families with BD and examined its associations with mood.

**Objective:** Using EMA, this study investigated the concurrent and predictive relationships between impulsivity and mood.

**Methods:** Multiplex family members with BD (n=8), unaffected family members (n=6), individuals with BD not from families (n=8) and healthy controls (n=8), completed daily EMA surveys about mood and impulsivity for 6-12 weeks. Mixed-effects regression concurrent and lagged models were employed to analyze the relationship between impulsivity and mood.

**Results:** The diagnosis of BD was associated with lower mood and higher impulsivity levels. Belonging to a multiplex family (i.e. high genetic load for BD) was associated with lower mood but not with higher impulsivity. BD participants showed a negative association between impulsivity and concurrent mood measured with EMA. Time-lagged analyses revealed a significant negative association between prior impulsivity and mood at the next assessment independent of diagnosis.

**Conclusions:** These results contribute to the understanding of the complex interactions between BD, the genetic load of the disorder, impulsivity and mood. Furthermore, these findings indicate the potential benefits of addressing impulsivity as a means to improve mood outcomes at an early stage.

## Introduction

Impulsivity is a frequently presenting component of bipolar disorder (BD) in its different phases and episodes, and has been proposed as a core feature of the disorder [1,2]. Furthermore, given the strong genetic load of BD, impairments of impulsivity have been observed in unaffected first-degree relatives [3,4]. Considering the important role of impulsivity in BD and its association with non adherence to medication, lower quality of life, higher functional disability, longer duration of illness and an increased number of suicide attempts [5–9], it is relevant to understand the relationship of impulsivity with different mood states.

Impulsivity is a complex and multidimensional construct without a widely agreed upon structure [10]. Several researchers have attempted to categorize it using different paradigms and have distinguished between: a) trait impulsivity and state impulsivity [11]; b) behavioural impulsivity versus self-reported impulsivity [10]; and c) impulsive choice and impulsive action [12]. Generally, impulsivity can be defined as a predisposition to rapid, unplanned reactions to internal or external stimuli that fail to take into account the negative consequences of those reactions to the individual themself or to others [13]. Its various dimensions have been commonly assessed with both self-report measures (e.g., the Barratt Impulsiveness Scale) [14] and behavioural measures (behavioural laboratory tasks). However, despite the enormous potential of these methods, none of them can capture the dynamic fluctuations of impulsivity.

BD involves mood fluctuations, sometimes very rapid, over a relatively short period of time. Therefore, to understand the disturbances that precede a mood change, it is particularly important to employ temporally sensitive methodologies that are not affected by retrospective biases. Recent years have highlighted the need for new types of ecologically valid measures that can provide a longitudinal assessment [15,16]. Psychiatric disorder symptoms can fluctuate rapidly, necessitating a more detailed and continuous assessment to identify critical transition points where timely intervention may be most effective. Modern Ecological Momentary Assessment (EMA) approaches, which longitudinally study individuals in their everyday natural environments using devices such as smartphones, hold promise for nuanced examination of the mechanisms of mood and impulsivity dysregulation [17,18].

Previous research using EMA approaches to explore mood and impulsivity in BD is limited, and findings are inconsistent. One pilot study compared BD patients with healthy controls (HC) on EMA measures of mood and impulsivity found no differences in mean impulsivity between the two groups, but higher variability in mean mood and impulsivity in the BD group compared to HC [19]. In another study, analysis of impulsivity measured by the EMA in BD patients found that negative, but not positive affect, predicted increases in impulsivity, which subsequently predicted decreases in positive affect [20]. Finally, Titone et al., 2022 found higher daily impulsivity in BD participants compared to HC participants. This same study demonstrated a bidirectional association between high impulsivity and high next-day negative affect and showed that impulsivity specifically predicted next-day anger and anxiety [21].

The present study investigated the concurrent and predictive relationships between impulsivity and mood in BD, using an EMA design with concurrent assessments.

## Methodology

### Setting and Participants

The study was conducted at the Regional University Hospital of Malaga (Spain) between 1 July 2020 and 30 November 2021. The participant sample consisted of a convenience selection of BD participants and unaffected healthy individuals (with at least one first-degree relative with an affective disorder) members of a cohort of families with high BD prevalence from the Andalusian Bipolar Family (ABiF) study [22,23], as well as age- and sex-matched BD participants and healthy controls from the general population. Inclusion and exclusion criteria are listed in Table 1.

**Table 1.** Inclusion/exclusion criteria.

| **Groups** | **Inclusion Criteria** | **Exclusion Criteria** |
| --- | --- | --- |
| All | – 18+ years old.<br>– fluently speak and read Spanish.<br>– familiar with smartphone operation. | – severe cognitive disabilities. |
| FP | – diagnosed with BD.<br>– member of a high density BD family.<br>– euthymic at the time of assessment. | – comorbid substance use disorder. |
| FC | – member of a high density BD family.<br>– first degree relative with affective disorder.<br>– no personal history of lifetime BD or MDD or psychotic disorder (schizophrenia, spectrum psychotic disorder). | – diagnosis of any affective disorder or substance use disorder. |
| P | – diagnosed with BD.<br>– not a member of one of the participating families.<br>– euthymic at the time of assessment.<br>– no first or second degree relatives with known history of BD or psychotic disorder (schizophrenia, spectrum psychotic disorder, MDD with psychotic symptoms). | – comorbid substance use disorder. |
| C | – not a member of one of the participating families.<br>– no personal history of lifetime BD or MDD or other psychotic.<br>– disorder (schizophrenia, spectrum psychotic disorder).<br>– no first or second degree relatives with known history of BD or other psychotic disorder (schizophrenia, spectrum psychotic disorder, MDD with psychotic symptoms). | – diagnosis of any affective disorder or substance use disorder. |

BD=Bipolar Disorder, MDD=Major Depressive Disorder, FP=Patients with BD who are members of multiplex BD families, FC= Unaffected members of multiplex BD families, P=Patients with BD from the general population, C=Healthy Controls

Thirty-four individuals were recruited to participate in the study. Four individuals dropped out before the end of the evaluation period, resulting in a final sample of 30 participants consisting of 14 males and 16 females, with a mean age of 52.60±14.42 years. Participants comprised 4 groups: 1) BD participants from families (BDF, n=8), family controls (FC, n=6), BD ambulatory cases (BDC, n=8), and healthy controls from the general population (HC, n=8). Diagnoses of BD were distributed as follows: BD type I (n=12; 5BDF,7BDC), BD type II (n=3; 2BDF,1BDC) and BD unspecified (n=1; 1BDF). At study entry, all participants were confirmed to be in a euthymic state.

The present study was carried out with the approval of the local ethics committee (Provincial Research Ethics Committee of Málaga), and written informed consent was obtained from all participants.

### Assessments and variables

The researchers provided participants with detailed explanations of the study and its procedure. First, inclusion and exclusion criteria were checked, after which sociodemographic data were collected and a structured general health questionnaire was administered. Subsequently, in a brief training session, participants received detailed instructions on the use of the movisensXS application (movisens GmbH, Karlsruhe, Germany, https://www.movisens.com/de/produkte/movisensxs), which was used for the assessment.

Mood and impulsivity were assessed using the movisensXS application implemented on the participants’ smartphones or those provided by the researchers (Samsung Galaxy J7). Participants completed the questionnaires three times a day (at 9:30 am, 3:30 pm and 8:30 pm) as instructed by the app. To measure mood, an item named ‘General mood’ was used with a visual analogue scale with a response option from 0 (downcast) to 100 (elevated). Impulsivity was assessed using The Momentary Impulsivity Scale, recommended for the EMA of the impulsivity construct [24]. Participants were asked to indicate the extent to which they felt this way in the last 15 minutes for each of the following items: 1) I said things without thinking; 2) I spent more money than I meant to; 3) I have felt impatient; 4) I made a “spur of the moment” decision. For each item, responses are selected on a Likert scale with the following options: 1: very little or not at all, 2: a little, 3: moderately, 4: quite a lot and 5: extremely. Data recorded in the movisensXS app on the smartphones were automatically uploaded to a secure server. Self-reported momentary assessments were made during a period of 6 to 12 weeks.

### Statistical analysis

Descriptive statistics were used to characterize the sample. For group comparisons, the Welch ANOVA test with a post-hoc Tukey test was used. Since longitudinal data with various measures over time were analyzed, mixed-effects regression models were used for analysis, with the individual as the random effect. To analyze the relationship between impulsivity and mood concurrent and lagged models were employed. We employed lagged models to assess how prior values of an independent variable predict subsequent changes in a dependent variable, enabling us to capture temporal dependencies and delayed effects within the longitudinal data. Two types of models were used: 1) one in which the dependent variable was mood and the independent variable was impulsivity, and 2) another in which the dependent variable was impulsivity and the independent variable was mood. Regarding the independent variables, two variables were introduced: 1) a time-invariant variable which was the individual’s mean throughout the evaluation period, and 2) a measure for each assessment (time-varying) that was centered around that person’s mean.

In the lagged model, the previous measure of the independent variable (t-1) was used as a predictor of the dependent variable’s value (t) in the subsequent momentary assessment. A model with random slopes and autoregressive correlation was used as it showed significantly better fit, as indicated by a lower Akaike Information Criterion. Age, gender, time, presence of bipolar diagnosis and belonging to a family with multiple cases of BD were included as variables in the regression models. Additionally, the interaction between the independent variable (time-varying) and BD diagnosis, as well as between the independent variable and belonging to families with multiple cases of BD, were included. Belonging to a family with multiple cases of BD was included as a variable to account for potential genetic or familial influences on the relationship between impulsivity and mood, as these factors may affect the severity, frequency, or pattern of these symptoms over time. The significance level was set at 0.0125 following Bonferroni correction by four comparisons (mood, impulsivity, diagnosis and belonging to families). R-Studio 2023.03.0+386 with R version 4.3.0. was used for the statistical analyses.

## Results

The sociodemographic and clinical characteristics of the sample, as well as the differences between groups, are shown in Table 2. There were significant differences in average mood between the groups (F=4.81, *p*=.017), with differences observed between BDF and HC (Diff =-31.70, *p*=.001) and BDC and HC (Diff=-25.94, *p*=.007), but no significant differences when compared with FC. No significant differences between groups in average impulsivity were found.

**Table 2.**
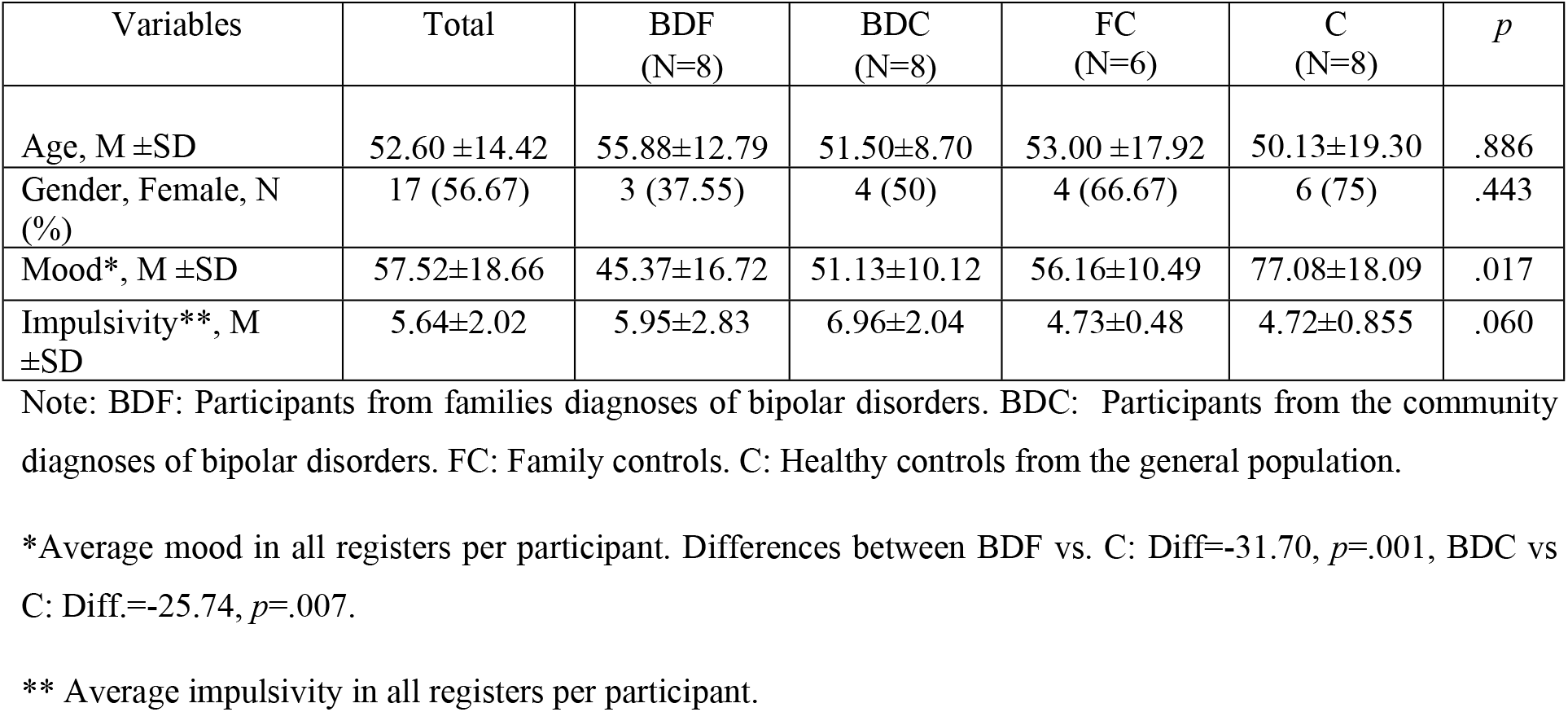
Sociodemographic and clinical characteristics of the sample.

### Mood as dependent variable

In the concurrent model, mean impulsivity during the assessment period was related to lower mood (B=-3.984, *p*=.007) and variation from the individual’s mean impulsivity was related to lower mood but in those diagnosed with BD as shown by the significant interaction results (B=-0.660, *p*=.002). In the lagged model, variation with respect to the individual’s mean impulsivity was related to lower mood (B=-0.596, *p*=.005) and in this case there was no interaction between impulsivity and diagnosis of BD (B=0.036, *p*=.863). More information on the conducted analyses is provide in Table 3.

**Table 3.** Concurrent and lagged mixed regression model using mood assessed by EMA as the dependent variable.

| Models (Dependent variable: Mood) | *Concurrent model* |  |  |  |  | *Lagged model (t-1)* |  |  |  |  |
| --- | --- | --- | --- | --- | --- | --- | --- | --- | --- | --- |
| **Fixed Effects** | *B* | *SE* | *Df* | *t* | *p* | *B* | *SE* | *Df* | *t* | *p* |
| Impulsivity (time varying) | 0.074 | 0.210 | 6733 | 0.350 | .726 | -0.596 | 0.210 | 6702 | -2.843 | .005 |
| Average Impulsivity (time invarying) | -3.984 | 1.338 | 24 | -2.977 | .007 | -3.957 | 1.347 | 24 | -2.938 | .007 |
| Bipolar diagnosis | -12.479 | 5.375 | 24 | -2.321 | .029 | -12.659 | 5.410 | 24 | -2.340 | .028 |
| Multiplex family belonging | -11.483 | 4.774 | 24 | -2.406 | .024 | -11.709 | 4.804 | 24 | -2.437 | .023 |
| Time | 0.011 | 0.016 | 6733 | -0.709 | .478 | -0.012 | 0.016 | 6702 | -0.750 | .453 |
| Age | -0.255 | 0.173 | 24 | -1.470 | .155 | -0.259 | 0.175 | 24 | -1.483 | .151 |
| Gender | -0.231 | 5.019 | 24 | -0.046 | .964 | -0.237 | 5.052 | 24 | -0.047 | .963 |
| ImpulsivityxFamily | -0.036 | 0.186 | 6733 | -0.195 | .845 | 0.335 | 0.185 | 6702 | 1.809 | .071 |
| ImpulsivityxDiagnosis | -0.660 | 0.211 | 6733 | -3.123 | .002 | 0.036 | 0.210 | 6702 | 0.173 | .863 |
| **Random effects** | AR1 | Intercept |  | Residuals |  | AR1 | Intercept |  | Residuals |  |
| Estimations | 0.372 | 14.857 |  | 12.712 |  | 0.376 | 14.961 |  | 12.684 |  |

### Impulsivity as dependent variable

Similar to the previous model, in the concurrent model, lower mean mood during the assessment period was related to higher impulsivity (B=-0.067, *p*=.002) and variation from the individual’s mean mood was related to impulsivity but in those diagnosed with BD as shown by the interaction results (B=-0.019, *p*<.001). However, in the lagged model, variation with respect to the individual’s mean mood was not related to subsequent change in impulsivity (B=-0.006, *p*=.107) and there was also no significant interaction with diagnosis of BD (B=0.004, *p*=.817). More information can be found in Table 4.

**Table 4.**
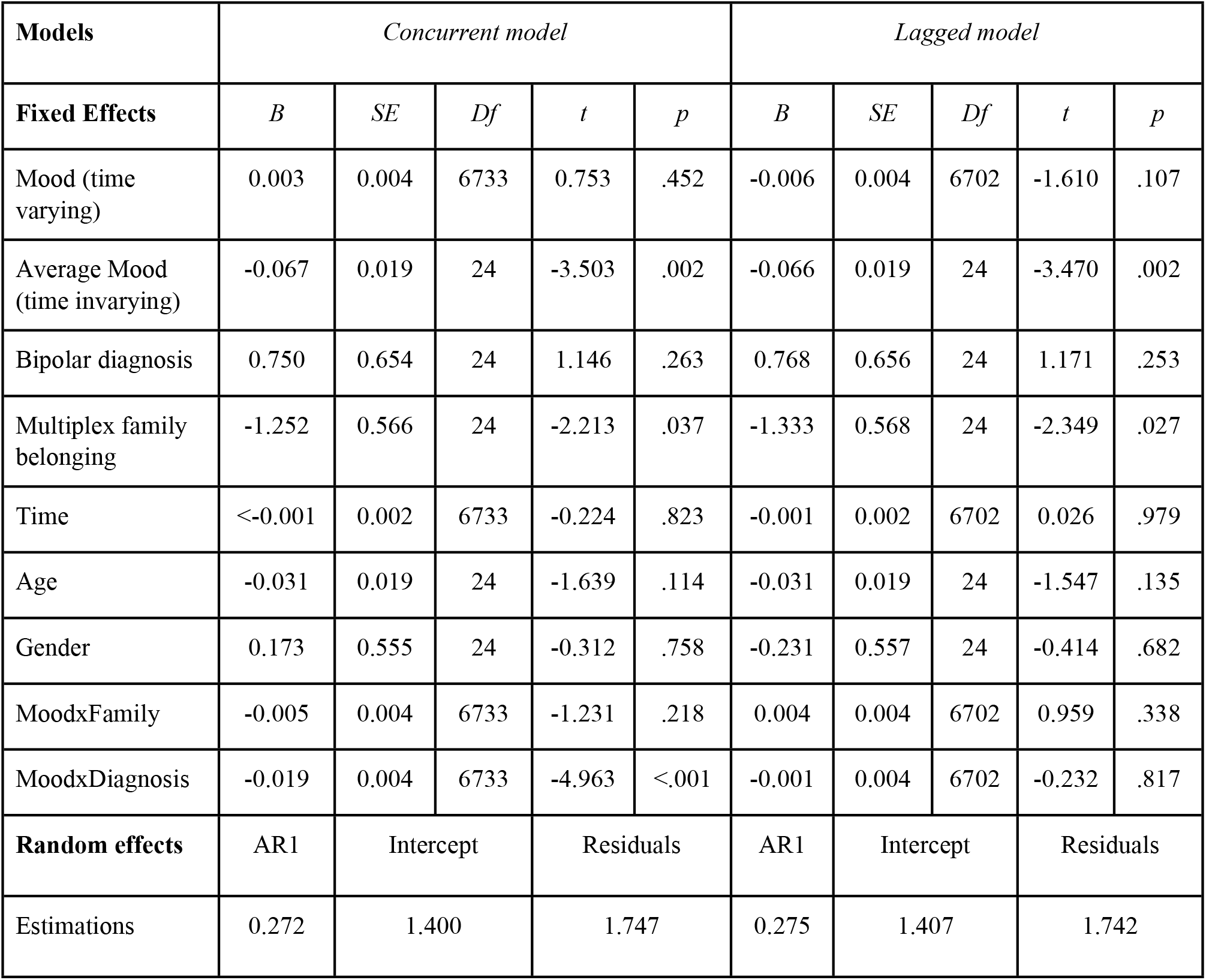
Concurrent and lagged mixed regression model using impulsivity as the dependent variable assessed by EMA.

| **Models** | *Concurrent model* |  |  |  |  | *Lagged model* |  |  |  |  |
| --- | --- | --- | --- | --- | --- | --- | --- | --- | --- | --- |
| **Fixed Effects** | *B* | *SE* | *Df* | *t* | *p* | *B* | *SE* | *Df* | *t* | *p* |
| Mood (time varying) | 0.003 | 0.004 | 6733 | 0.753 | .452 | -0.006 | 0.004 | 6702 | -1.610 | .107 |
| Average Mood (time invarying) | -0.067 | 0.019 | 24 | -3.503 | .002 | -0.066 | 0.019 | 24 | -3.470 | .002 |
| Bipolar diagnosis | 0.750 | 0.654 | 24 | 1.146 | .263 | 0.768 | 0.656 | 24 | 1.171 | .253 |
| Multiplex family belonging | -1.252 | 0.566 | 24 | -2.213 | .037 | -1.333 | 0.568 | 24 | -2.349 | .027 |
| Time | <-0.001 | 0.002 | 6733 | -0.224 | .823 | -0.001 | 0.002 | 6702 | 0.026 | .979 |
| Age | -0.031 | 0.019 | 24 | -1.639 | .114 | -0.031 | 0.019 | 24 | -1.547 | .135 |
| Gender | 0.173 | 0.555 | 24 | -0.312 | .758 | -0.231 | 0.557 | 24 | -0.414 | .682 |
| MoodxFamily | -0.005 | 0.004 | 6733 | -1.231 | .218 | 0.004 | 0.004 | 6702 | 0.959 | .338 |
| MoodxDiagnosis | -0.019 | 0.004 | 6733 | -4.963 | <.001 | -0.001 | 0.004 | 6702 | -0.232 | .817 |
| **Random effects** | AR1 | Intercept |  | Residuals |  | AR1 | Intercept |  | Residuals |  |
| Estimations | 0.272 | 1.400 |  | 1.747 |  | 0.275 | 1.407 |  | 1.742 |  |

## Discussion

The study found a significant negative association between impulsivity and concurrent mood at the EMA in those with BD. Furthermore, a significant relationship was found between prior impulsivity and mood at the next assessment independent of diagnosis, whereas the reverse did not occur, no significant association was found between mood at the EMA and impulsivity at the subsequent assessment. These results support the hypothesis that impulsivity could be a risk factor for subsequent downcast mood, although further studies are needed to confirm this hypothesis. Additionally, those with BD had a downcast mood in comparison with controls but not with controls from the families.

There is a large body of evidence showing that impulsivity plays a role in BD [1,25]. The present study also found that BD play a moderator role in the relationship between impulsivity and mood. The results of the study regarding the association of BD with negative mood reported using daily monitoring are in agreement with the study of Schwartz et al. (2016). Other studies have found higher trait and behavioural impulsivity in relatives of patients diagnosed with BD [26]. However, in this study we found a non-significant relationship between belonging a multiple family and impulsivity. This result could be influenced by the small size of the study groups.

There is a growing body of evidence linking impulsivity and negative mood [27–29] in both non clinical [30,31] and clinical populations [32]. One hypotheses proposed to explain this relationship is that both conditions share a common serotonergic pathway [33]. This relationship has also been found when studies have been conducted using ecological intensive measurement. In our study, the presence of BD was found to moderate the relationship between impulsivity and mood at the same time point. Regarding the temporal relationships between these two variables, the study by Deep et al., 2016 found bidirectional relationships between impulsivity and affect measured with EMA in a sample of participants with BD [20]. Similarly, in the study by Titone et al., 2022 a bidirectional relationship between impulsivity and mood was found [34]. However, in the present study, in the lagged models, there was only a significant relationship between impulsivity and affect in one direction. There may be different pathways through which impulsivity affects mood. For example, there is a growing body of evidence linking impulsivity to a lower likelihood of positive events, decreased functioning [35] and poorer sleep quality [21,36].

## Limitations

To interpret these results, the limitations of the study must be considered. The main limitation is the small sample size even though a long and intensive assessment was carried out. Also, the EMA measure of impulsivity is based on four questions that may not reflect the construct in its entirety and should be considered preliminary as it has not been validated. Similarly, mood was measured with a single question that may not reflect the complexity of mood.

## Conclusion

Our results indicate that based in intensive longitudinal data a relationship between impulsivity and concurrent negative mood in those with BD and a relationship between previous impulsivity and negative mood at the next assessment regardless of diagnosis were found. These results contribute to understanding the complex interactions between BD, the genetic load of the disorder, impulsivity and mood. Our findings suggest that impulsivity could be a potential target for improving mood in individuals with bipolar disorder; however, further research with larger samples is needed to better understand this relationship and its potential to guide effective interventions.

## Acknowledgments

We would like to thank all the participants for their commitment to and participation in the study.

## Conflict of interest

All authors declare that they have no conflict of interest.

### Abbreviations

AbiF: Andalusian Bipolar Family study
ANOVA: Analysis of Variance
BD: Bipolar disorder
BDC: Bipolar disorder ambulatory cases
BDF: Bipolar disorder participants from families
EMA: Ecological Momentary Assessment
FC: family controls

## Data Availability

The datasets are available from the corresponding author on reasonable request

## References

1. Ramírez-Martín A, Ramos-Martín J, Mayoral-Cleries F, Moreno-Küstner B, Guzman-Parra J. Impulsivity, decision-making and risk-taking behaviour in bipolar disorder: a systematic review and meta-analysis. Psychol Med. 2020; 1–13. doi:10.1017/S0033291720003086

2. Strakowski SM, Fleck DE, DelBello MP, Adler CM, Shear PK, Kotwal R, et al. Impulsivity across the course of bipolar disorder. Bipolar Disord. 2010;12:285–297. doi:10.1111/j.1399-5618.2010.00806.x

3. Lombardo LE, Bearden CE, Barrett J, Brumbaugh MS, Pittman B, Frangou S, et al. Trait impulsivity as an endophenotype for bipolar I disorder. Bipolar Disord. 2012;14:565. doi:10.1111/J.1399-5618.2012.01035.X

4. Sanches M, Scott-Gurnell K, Patel A, Caetano SC, Zunta-Soares GB, Hatch JP, et al. Impulsivity in Children and Adolescents with Mood Disorders and Unaffected Offspring of Bipolar parents. Compr Psychiatry. 2014. doi:10.1016/j.comppsych.2014.04.018

5. Ekinci O, Albayrak Y, Ekinci AE, Caykoylu A. Relationship of trait impulsivity with clinical presentation in euthymic bipolar disorder patients. Psychiatry Res. 2011;190:259–264. doi:10.1016/j.psychres.2011.06.010

6. Jiménez E, Arias B, Castellví P, Goikolea JM, Rosa AR, Fañanás L, et al. Impulsivity and functional impairment in bipolar disorder. J Affect Disord. 2012;136:491–497. doi:10.1016/j.jad.2011.10.044

7. Victor SE, Johnson SL, Gotlib IH. Quality of life and impulsivity in bipolar disorder. Bipolar Disord. 2011;13:303–309. doi:10.1111/j.1399-5618.2011.00919.x

8. Belzeaux R, Boyer L, Mazzola-Pomietto P, Michel P, Correard N, Aubin V, et al. Adherence to medication is associated with non-planning impulsivity in euthymic bipolar disorder patients. J Affect Disord. 2015;184. doi:10.1016/j.jad.2015.05.041

9. Kulacaoglu F, Izci F. The Effect of Emotional Dysregulation and Impulsivity on Suicidality in Patients with Bipolar DisorderER. Psychiatr Danub. 2022;33:706–714. doi:10.24869/PSYD.2022.706

10. Sharma L, Kohl K, Morgan TA, Clark LA. “Impulsivity”: Relations between self-report and behavior. J Pers Soc Psychol. 2013;104:559–575. doi:10.1037/a0031181

11. Halvorson MA, Pedersen SL, Feil MC, Lengua LJ, Molina BSG, King KM. Impulsive States and Impulsive Traits: A Study of the Multilevel Structure and Validity of a Multifaceted Measure of Impulsive States. Assessment. 2021;28:796–812. doi:10.1177/1073191120939161

12. McCarthy DE, Minami H, Bold KW, Yeh VM, Chapman G. Momentary assessment of impulsive choice and impulsive action: Reliability, stability, and correlates. Addict Behav. 2018;83:130–135. doi:10.1016/j.addbeh.2017.11.031

13. Moeller FG, Barratt ES, Dougherty DM, Schmitz JM, Swann AC. Psychiatric aspects of impulsivity. Am J Psychiatry. 2001;158:1783–1793. doi:10.1176/appi.ajp.158.11.1783

14. Patton Jim H, Stanford Matthew SBES. Factor structure of the barratt impulsiveness scale. Journal of Clinical Psychology. 1995. pp. 768–774.

15. Keramatian K, Torres IJ, Yatham LN. Neurocognitive functioning in bipolar disorder: What we know and what we don’t. Dialogues Clin Neurosci. 2022;23:29–38. doi:10.1080/19585969.2022.2042164

16. Wenzel M, Kubiak T, Ebner-Priemer UW. Ambulatory assessment as a means of longitudinal phenotypes characterization in psychiatric disorders. Neurosci Res. 2016;102:13–21. doi:10.1016/j.neures.2014.10.018

17. Trull TJ, Ebner-Priemer U. Ambulatory assessment. Annual Review of Clinical Psychology. 2013. pp. 151–176. doi:10.1146/annurev-clinpsy-050212-185510

18. Shiffman S, Stone AA, Hufford MR. Ecological momentary assessment. Annu Rev Clin Psychol. 2008;4:1–32.

19. Schwartz S, Schultz S, Reider A, Saunders EFH. Daily mood monitoring of symptoms using smartphones in bipolar disorder: A pilot study assessing the feasibility of ecological momentary assessment. J Affect Disord. 2016;191:88–93. doi:10.1016/j.jad.2015.11.013

20. Depp CA, Moore RC, Dev SI, Mausbach BT, Eyler LT, Granholm EL. The temporal course and clinical correlates of subjective impulsivity in bipolar disorder as revealed through ecological momentary assessment. J Affect Disord. 2016;193:145–150. doi:10.1016/j.jad.2015.12.016

21. Titone MK, Goel N, Ng TH, MacMullen LE, Alloy LB. Impulsivity and sleep and circadian rhythm disturbance predict next-day mood symptoms in a sample at high risk for or with recent-onset bipolar spectrum disorder: An ecological momentary assessment study. J Affect Disord. 2022;298:17–25. doi:10.1016/j.jad.2021.08.155

22. Guzman-Parra J, Rivas F, Strohmaier J, Forstner A, Streit F, Auburger G, et al. The Andalusian Bipolar Family (ABiF) Study: Protocol and sample description. Rev Psiquiatr Salud Ment. 2018;11:199–207. doi:10.1016/j.rpsm.2017.03.004

23. Guzman-Parra J, Streit F, Forstner AJ, Strohmaier J, González MJ, Gil Flores S, et al. Clinical and genetic differences between bipolar disorder type 1 and 2 in multiplex families. Transl Psychiatry. 2021;11. doi:10.1038/s41398-020-01146-0

24. Tomko RL, Solhan MB, Carpenter RW, Brown WC, Jahng S, Wood PK, et al. Measuring impulsivity in daily life: The momentary impulsivity scale. Psychol Assess. 2014;26:339–349. doi:10.1037/a0035083

25. Ramírez-Martín A, Sirignano L, Streit F, Foo JC, Forstner AJ, Frank J, et al. Impulsivity, decision-making, and risk behavior in bipolar disorder and major depression from bipolar multiplex families. Brain Behav. 2024;14. doi:10.1002/brb3.3337

26. Powers RL, Russo M, Mahon K, Brand J, Braga RJ, Malhotra AK, et al. Impulsivity in bipolar disorder: relationships with neurocognitive dysfunction and substance use history. Bipolar Disord. 2013;15:876–84. doi:10.1111/bdi.12124

27. Sperry SH, Lynam DR, Kwapil TR. The convergence and divergence of impulsivity facets in daily life. J Pers. 2018;86:841–852. doi:10.1111/JOPY.12359

28. Law MK, Fleeson W, Arnold EM, Michael Furr R. Using negative emotions to trace the experience of borderline personality pathology: Interconnected relationships revealed in an experience sampling study. J Pers Disord. 2016;30:52. doi:10.1521/PEDI_2015_29_180

29. Tomko RL, Lane SP, Pronove LM, Treloar HR, Brown WC, Solhan MB, et al. Undifferentiated Negative Affect and Impulsivity in Borderline Personality and Depressive Disorders: A Momentary Perspective. J Abnorm Psychol. 2015;124:740. doi:10.1037/ABN0000064

30. Schmidt RE, Gay P, Ghisletta P, Van Der Linden M. Linking impulsivity to dysfunctional thought control and insomnia: a structural equation model. J Sleep Res. 2010;19:3–11. doi:10.1111/J.1365-2869.2009.00741.X

31. Schmidt RE, Van der Linden M. The Aftermath of Rash Action: Sleep-Interfering Counterfactual Thoughts and Emotions. Emotion. 2009;9:549–553. doi:10.1037/A0015856

32. Rosen PJ, Factor PI. Emotional Impulsivity and Emotional and Behavioral Difficulties Among Children With ADHD. http://dx.doi.org/101177/1087054712463064. 2012;19:p779–793. doi:10.1177/1087054712463064

33. Apter A, van Praag HM, Plutchik R, Sevy S, Korn M, Brown SL. Interrelationships among anxiety, aggression, impulsivity, and mood: A serotonergically linked cluster? Psychiatry Res. 1990;32:191–199. doi:10.1016/0165-1781(90)90086-K

34. Titone MK, Depp C, Klaus F, Carrasco J, Young JW, Eyler LT. The interplay of daily affect and impulsivity measured by mobile surveys in bipolar disorder. Int J Bipolar Disord. 2022;10:25. doi:10.1186/S40345-022-00270-8

35. Walerius DM, Reyes RA, Rosen PJ, Factor PI. Functional Impairment Variability in Children With ADHD Due to Emotional Impulsivity. https://doi.org/101177/1087054714561859. 2014;22:724–737. doi:10.1177/1087054714561859

36. Patapoff M, Ramsey M, Titone M, Kaufmann CN, Malhotra A, Ancoli-Israel S, et al. Temporal relationships of ecological momentary mood and actigraphy-based sleep measures in bipolar disorder. J Psychiatr Res. 2022;150:257. doi:10.1016/J.JPSYCHIRES.2022.03.055

